# Long-COVID in Children and Adolescents: A Systematic Review and Meta-analyses

**DOI:** 10.1101/2022.03.10.22272237

**Authors:** Sandra Lopez-Leon, Talia Wegman-Ostrosky, Norma Cipatli Ayuzo del Valle, Carol Perelman, Rosalinda Sepulveda, Paulina A Rebolledo, Angelica Cuapio, Sonia Villapol

## Abstract

The objective of this systematic review and meta-analyses is to estimate the prevalence of long-COVID in children and adolescents and to present the full spectrum of symptoms present after acute COVID-19. We have used PubMed and Embase to identify observational studies published before February 10th, 2022 that included a minimum of 30 patients with ages ranging from 0 to 18 years that met the National Institute for Healthcare Excellence (NICE) definition of long-COVID, which consists of both ongoing (4 to 12 weeks) and post-COVID-19 (≥12 weeks) symptoms. Random-effects meta-analyses were performed using the MetaXL software to estimate the pooled prevalence with a 95% confidence interval (CI). Heterogeneity was assessed using I^2^ statistics. The Preferred Reporting Items for Systematic Reviewers and Meta-analysis (PRISMA) reporting guideline was followed (registration PROSPERO CRD42021275408). The literature search yielded 8,373 publications, of which 21 studies met the inclusion criteria, and a total of 80,071 children and adolescents were included. The prevalence of long-COVID was 25.24%, and the most prevalent clinical manifestations were mood symptoms (16.50%), fatigue (9.66%), and sleep disorders (8.42%). Children infected by SARS-CoV-2 had a higher risk of persistent dyspnea, anosmia/ageusia, and/or fever compared to controls. Limitations of the studies analyzed include lack of standardized definitions, recall, selection, misclassification, nonresponse and/or loss of follow-up, and a high level of heterogeneity.

## INTRODUCTION

It has been two years since the coronavirus disease 2019 (COVID-19) pandemic was first declared. Consequently, millions of cases and thousands of deaths have been reported worldwide^1^. Still, treatments have been developed rapidly during this time, and effective vaccines have been widely administered to the population, both children and adults, protecting millions from severe disease and death ^2^. Until now, the focus was primarily aimed at the acute phase of the disease. However, many individuals experience debilitating COVID-19 symptoms months later, requiring additional medical attention and follow-up.

Severe COVID-19 is less common in children than in adults^3^; however, two long-term consequences occur following severe acute respiratory syndrome coronavirus 2 (SARS-CoV-2) infection in children: multisystem inflammatory syndrome (MIS-C) and long-COVID. Both consequences can even appear in asymptomatic patients^4^. MIS-C is a condition where different body parts become inflamed^4^, it occurs in less than 0.01% of children infected and requires intensive care support in 68% of cases^5^. Long-COVID is a heterogeneous multisystemic condition for which there is still no precise definition and includes signs and symptoms that persist, develop, or fluctuate after SARS-CoV-2 infection. In October 2021, the WHO proposed a clinical definition for post-COVID-19 through a Delphi consensus stating it generally occurs three months from the onset of COVID-19, with symptoms lasting at least two months and cannot be explained by an alternative diagnosis^6^. On February 2nd, 2022, the National Institute for Health and Care Excellence (NICE) published a guideline defining long-COVID as signs and symptoms that continue or develop after acute COVID-19. This includes ongoing symptomatic COVID-19 (from 4 to 12 weeks) and post-COVID-19 syndrome (12 weeks or more)^7^. Other organizations, such as the National Institutes of Health (N.I.H.), also define long-COVID as post-acute symptoms after four weeks^8^. Many authors have used the following terms interchangeably when referring to long-COVID: long-haulers, COVID-long, post-acute sequelae of COVID-19 (PASC), post-COVID, COVID syndrome, and long-COVID. In the present study, we will use the generic definition from NICE and N.I.H., referring to long-COVID.

To date, most of the published research on long-COVID primarily focuses on adult populations, and there is limited information on pediatric populations^9,10^. The most recent meta-analysis has studied the long-COVID symptoms and their prevalence, risk factors, type, and duration, including studies up to July 2021, encompassing 23,141 children and young people^9^. The most common symptoms were fatigue 47%, dyspnea 43%, and headache 35%. In addition, compared to controls, the prevalence of cognitive difficulties, headache, loss of smell, sore throat, and sore eyes was statistically higher^9^, however, due to the lack of data, this study could only compute the pooled prevalence for 10 symptoms. To date, the potential range of signs and symptoms and their frequency of occurrence in children and adolescents remains unclear^11^. There is a need to create awareness among parents, physicians, and researchers on the afflictions following COVID-19 infection and the health system to better understand the sequelae to provide targeted medical attention and treatment. This systematic review and meta-analyses aim to estimate the prevalence of long-COVID in children and adolescents and identify the full spectrum of symptoms.

## METHODS

### Data sources and search strategy

This systematic review and meta-analyses were reported following the Preferred Reporting Items for Systematic Reviews and Meta-analyses (PRISMA) reporting guideline^12^. It examines the prevalence of long-COVID signs and symptoms in children under the age of 18 with a diagnosed case of COVID-19 (confirmed via real-time reverse transcription-polymerase chain reaction (rt-PCR), antigen or antibody (or serology) tests). To achieve this, two independent investigators (T.W.O and S.L.L.) searched PubMed and Embase to identify studies that met the following criteria: 1) a minimum of 30 patients, 2) ages ranged from 0 to 18 years, 3) studies published in English, 4) published before February 10th, 2022, and 5) meet the NICE definition of long-COVID, 6) excluding cohorts of children composed of exclusively pre-existing chronic diseases, or exclusively of MIS-C in children, and 7) excluding references of editorials, reviews, and commentaries (Figure 1).

**Figure 1.**
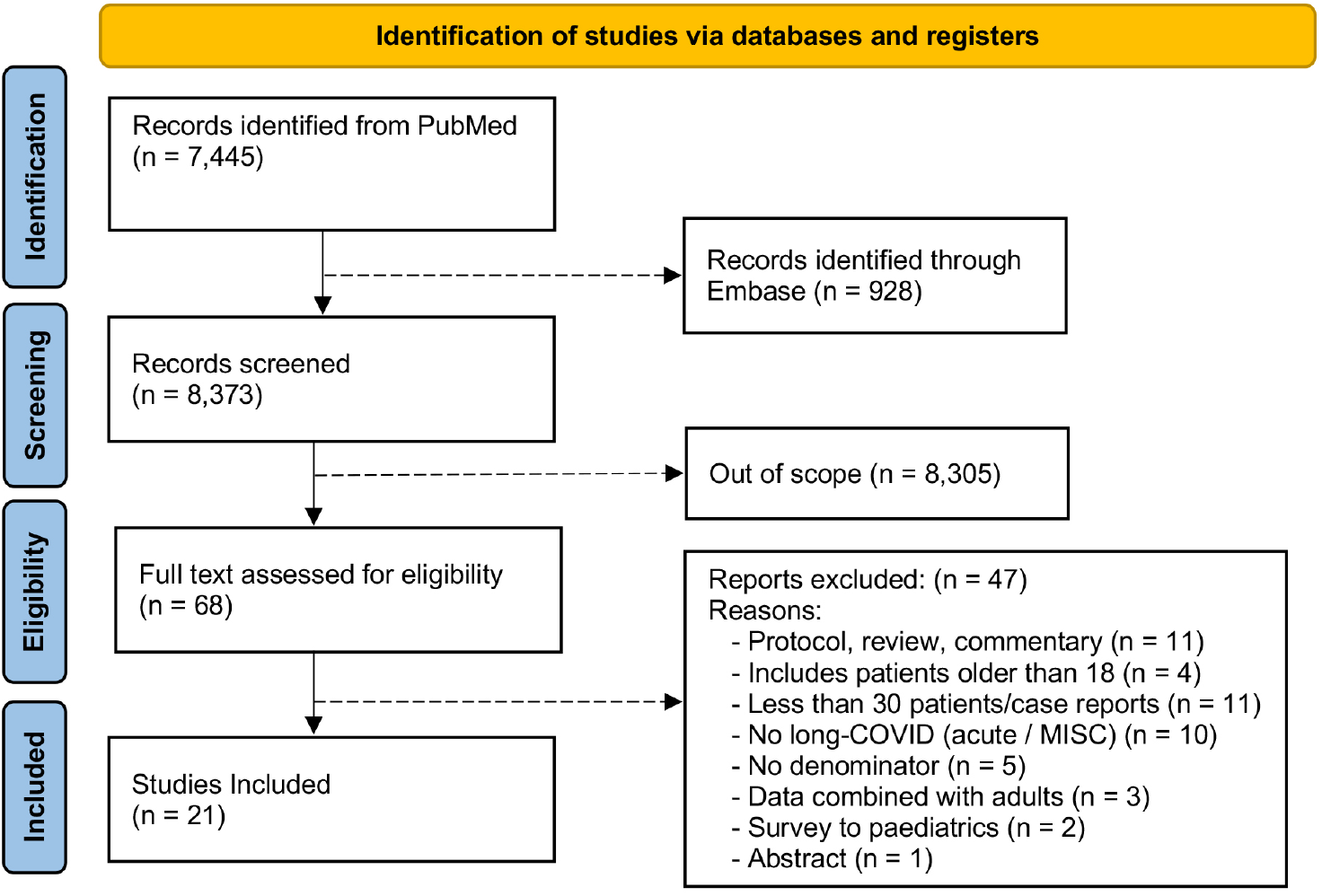
PRISMA diagram with exclusion criteria. Preferred Items for Systematic Reviews and Meta-Analyses (PRISMA) screening process flow. Out of 8,373 identified studies and after applying the inclusion and exclusion criteria, 21 studies were included in the quantitative synthesis.

The search terms used to identify publications discussing long-COVID in children were: (COVID-19 OR COVID OR SARSCOV-2 OR coronavirus OR “long-COVID” OR “post COVID”) AND (PASC OR haulers OR lingering OR “post-acute” OR persistent OR convalescent OR convalescence OR sequelae OR post-viral) AND (pediatric OR kids OR young OR infant OR children OR adolescents). Given that MedLine was included in the PubMed search, we excluded articles from MedLine in the Embase search along with those not related to COVID-19. Observational studies, including cohorts and cross-sectional studies, were analyzed only when the cases (numerator) were part of a COVID-19 cohort (denominator). Titles, abstracts, and full texts of articles were independently screened. Disagreement on including a full-text article was discussed among all the authors. We developed and registered a review protocol (PROSPERO registration number: CRD42021275408).

### Screening and data extraction

Data were extracted by four authors (A.C., C.A., P.R., R.S.) and Quality-Controlled (QCed) data by two authors (T.W.O., C.P.). Discrepancies were discussed with a third author. The descriptive variables extracted were country, study design, period of study, collection mode, follow-up time, the severity of COVID-19, sample size, COVID-19 diagnosis, age, percentage of males, outcomes, and names used to describe the long-term effects of COVID-19.

### Statistical analysis

Random-effects meta-analyses were performed for symptoms reported in two or more studies using MetaXL software to estimate the pooled prevalence, which uses a double arcsine transformation^13^. Prevalence (presented as percentages) with 95% confidence intervals (C.I.s) was estimated. Numerators represented the number of children with long-COVID, and denominators described the total number of children with acute COVID-19 (with and without long-term effects). To compare cases and controls adjusted for confounders, we used Review Manager (RevMan) software 5.4 to estimate the Odds Ratios (O.R.s)^14^. A p-value < 0.05 was considered statistically significant. Given the heterogeneity expected, a random-effects model was employed using the I^2^ statistics. Values of 25, 50, and 75% for I^2^ represented low, medium, and high heterogeneity, respectively. Studies with high precision plotted near the average, and those with low precision are speeded evenly on both sides of the average. Deviation from a funnel shape distribution this shape can indicate publication bias^14^. The quality control of the study was assessed using the QCed data. The MetaXL Guidelines describe and recommend this index, which evaluates the quality of studies assessing prevalence.

## RESULTS

### General characteristics of studies

The title and abstract of 8,373 publications were screened. After duplicates were removed, the search identified 68 publications after screening titles and abstracts, and 47 were excluded because they did not fulfill the inclusion criteria. A total of 21 studies were selected for the analyses (Figure 1). The general study characteristics are shown in Table 1. Different authors have used the terms “Post-acute COVID”, “long-COVID,” “Persistent COVID,” and “Persistent COVID Symptoms” as synonyms. Most of the studies assessed pre-specified symptoms included in a questionnaire. 18 of the selected studies come from European cohorts (Denmark, Russia, Italy, Germany, Tukey, Latvia, UK, France, Sweden, and Switzerland), one from Iran, one from Brazil, and one from Australia. The studies by Kikkenborg^15^ and Borch^16^ included an overlapping population (Denmark), as did Roge^17^ and Smane^18^ (Latvia). To ensure that no overlapping data were included, only the study with the largest sample size was included to estimate long-COVID and for outcomes reported in both studies. Still, several outcomes were only presented in one of the studies, therefore both studies were included in the overall meta-analysis. Four studies included only hospitalized patients, and the rest included all COVID-19 severities (asymptomatic, mild, moderate, and severe). The number of patients included in the studies ranged from 53 to 57,763, and ages ranged from 0 to 18 years. A total of 80,071 children and adolescents with COVID-19 were included in the meta-analyses. We identified more than 40 long-term clinical manifestations associated with COVID-19 in the literature reviewed (Table 2).

**Table 1.**
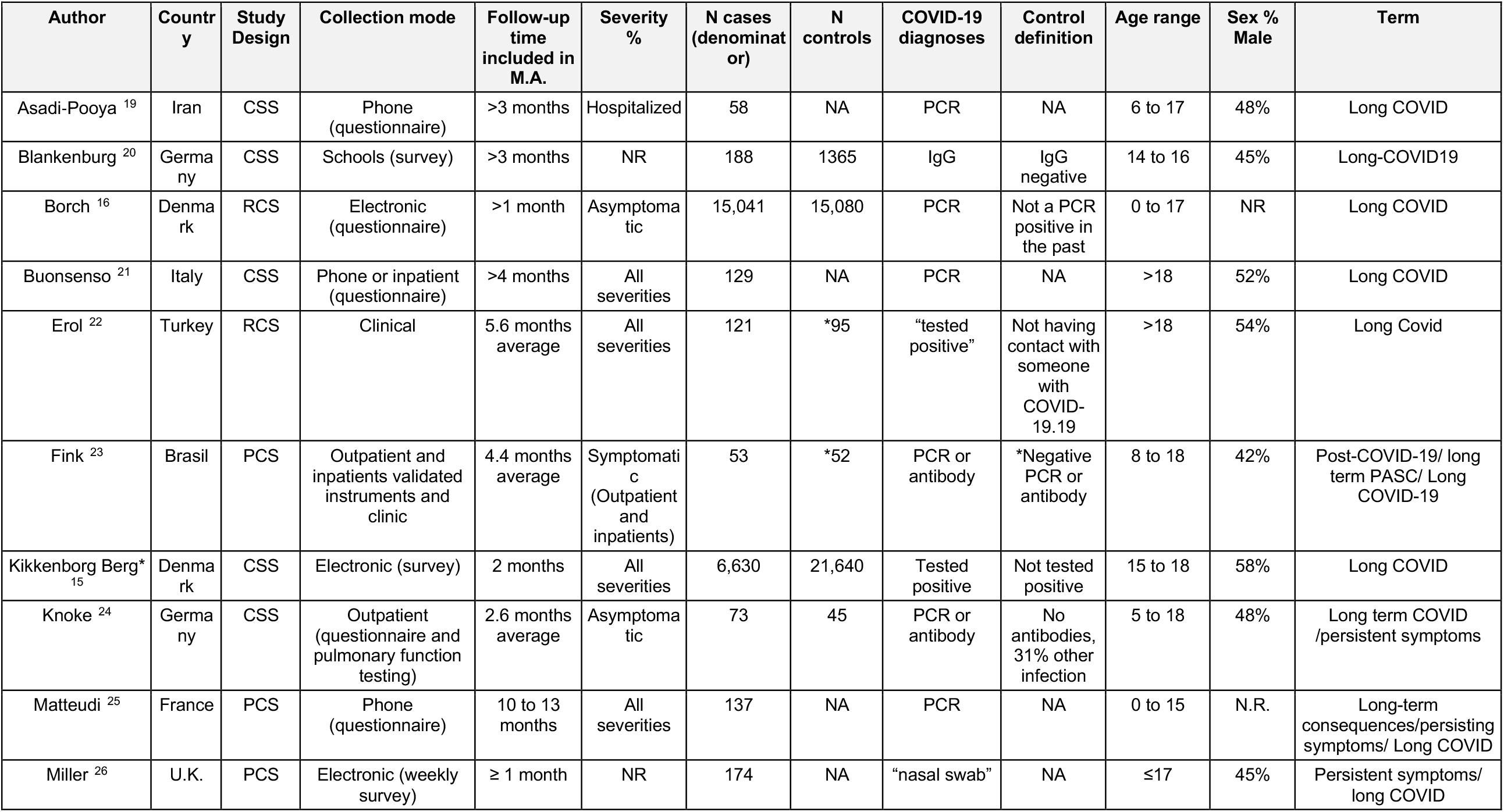
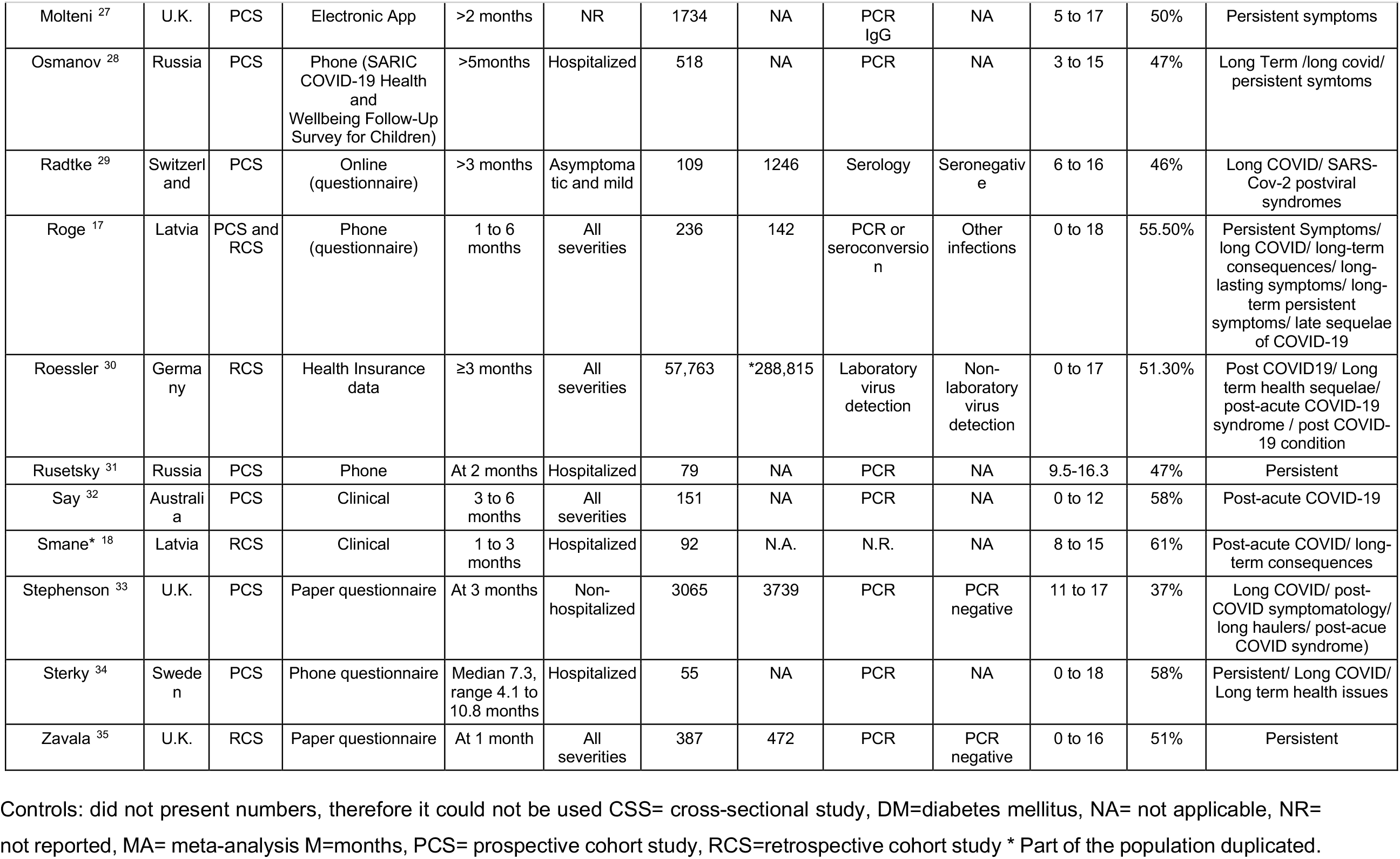
General Characteristics of Studies.

**Table 2.**
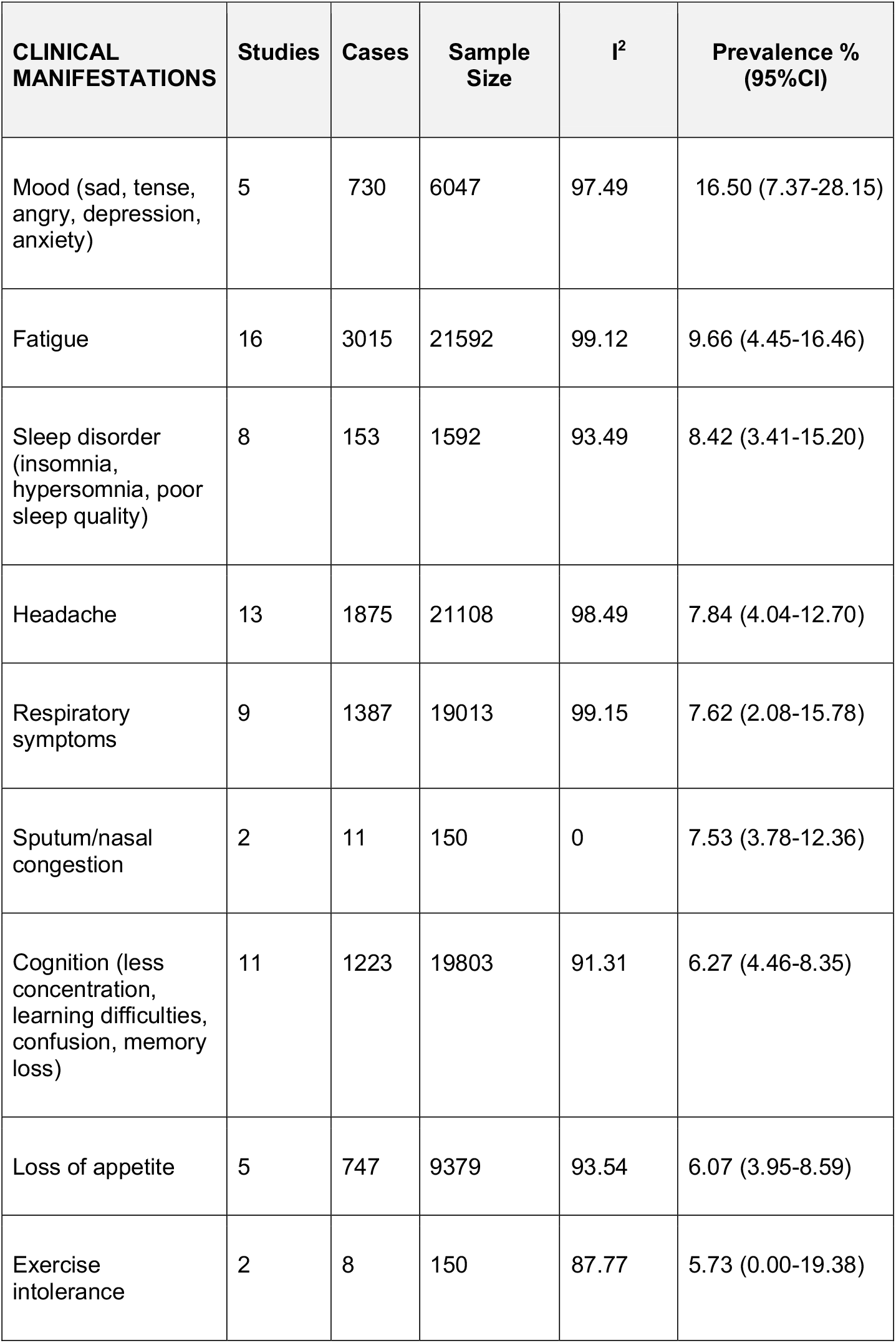
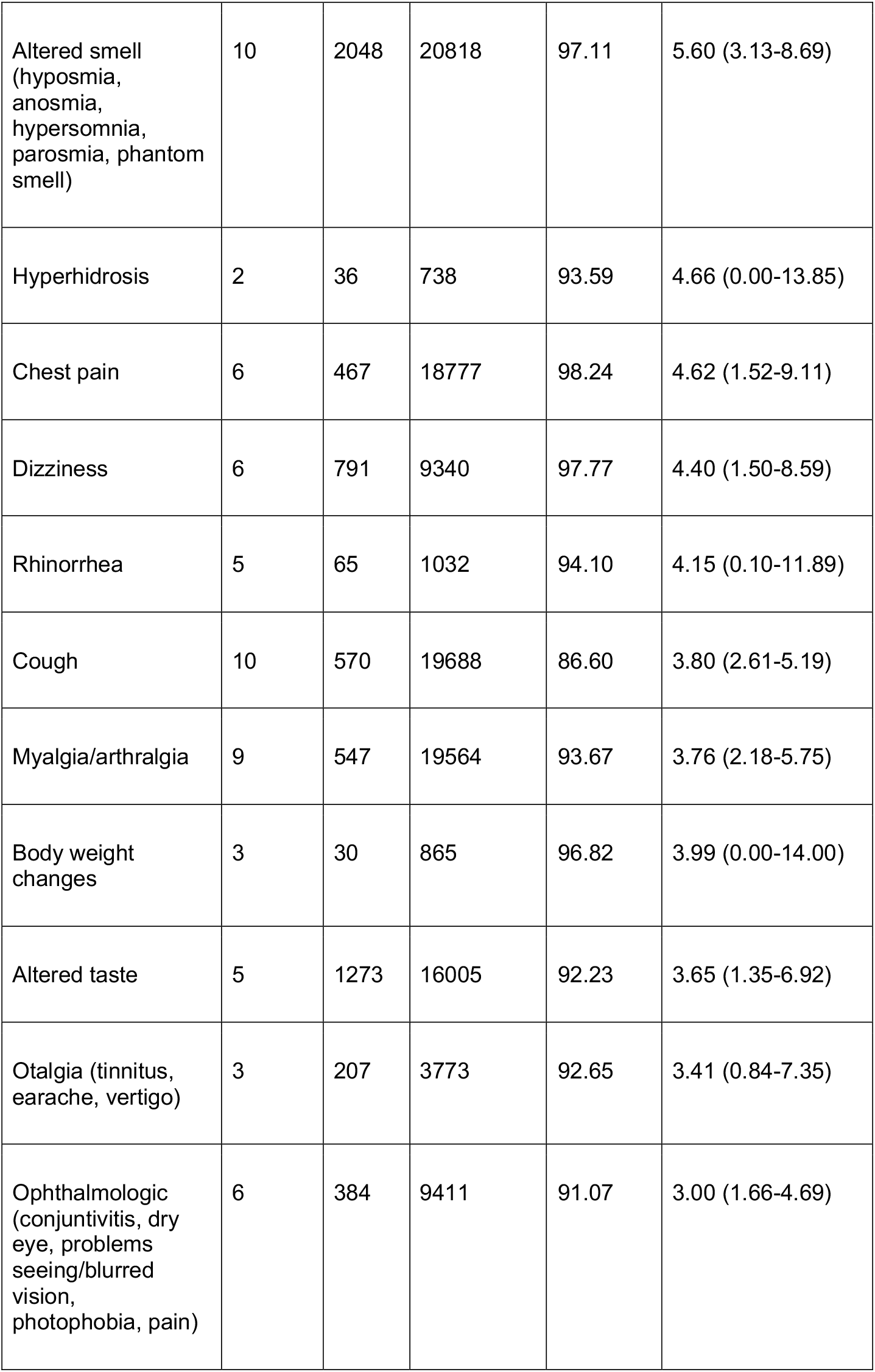
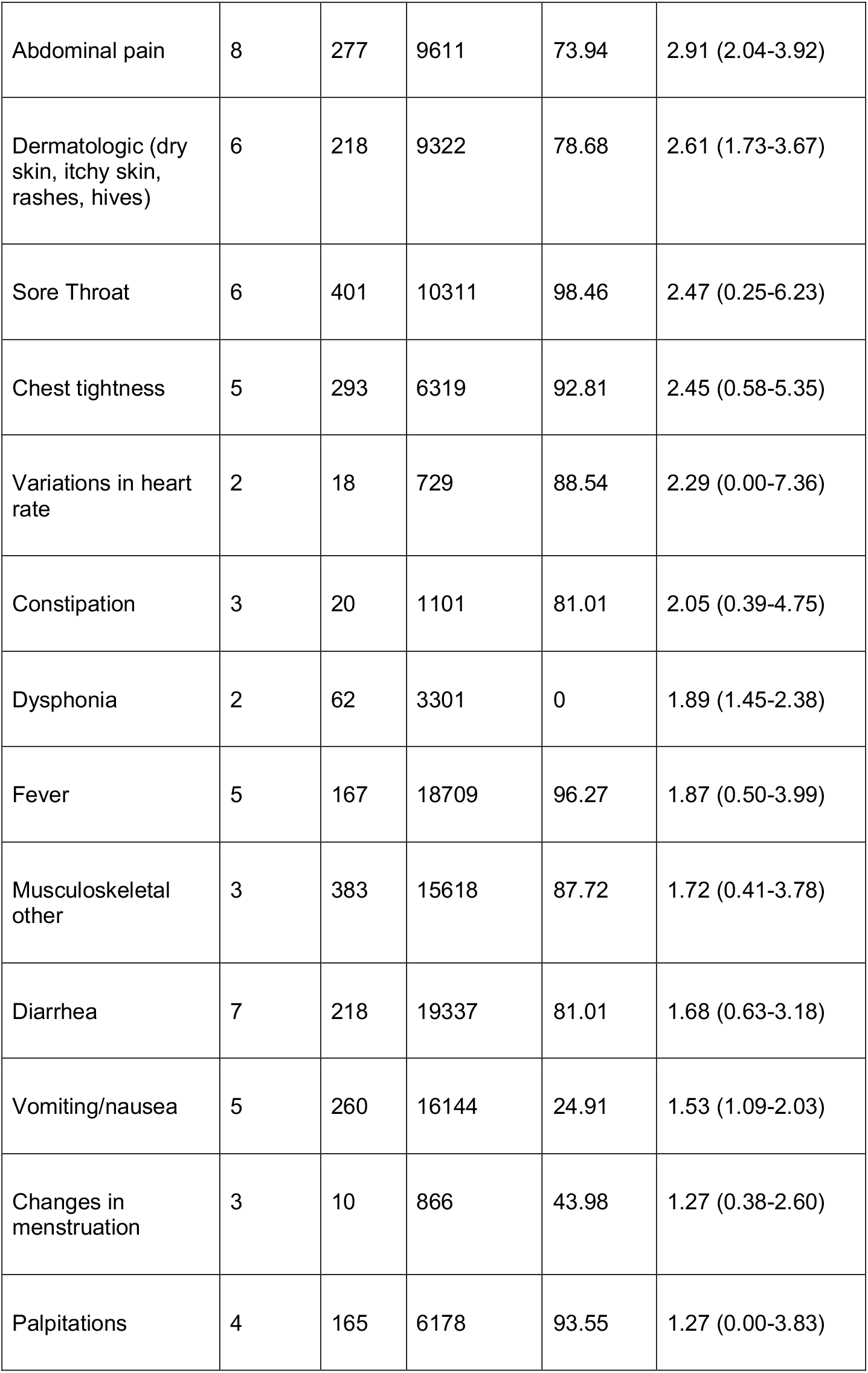
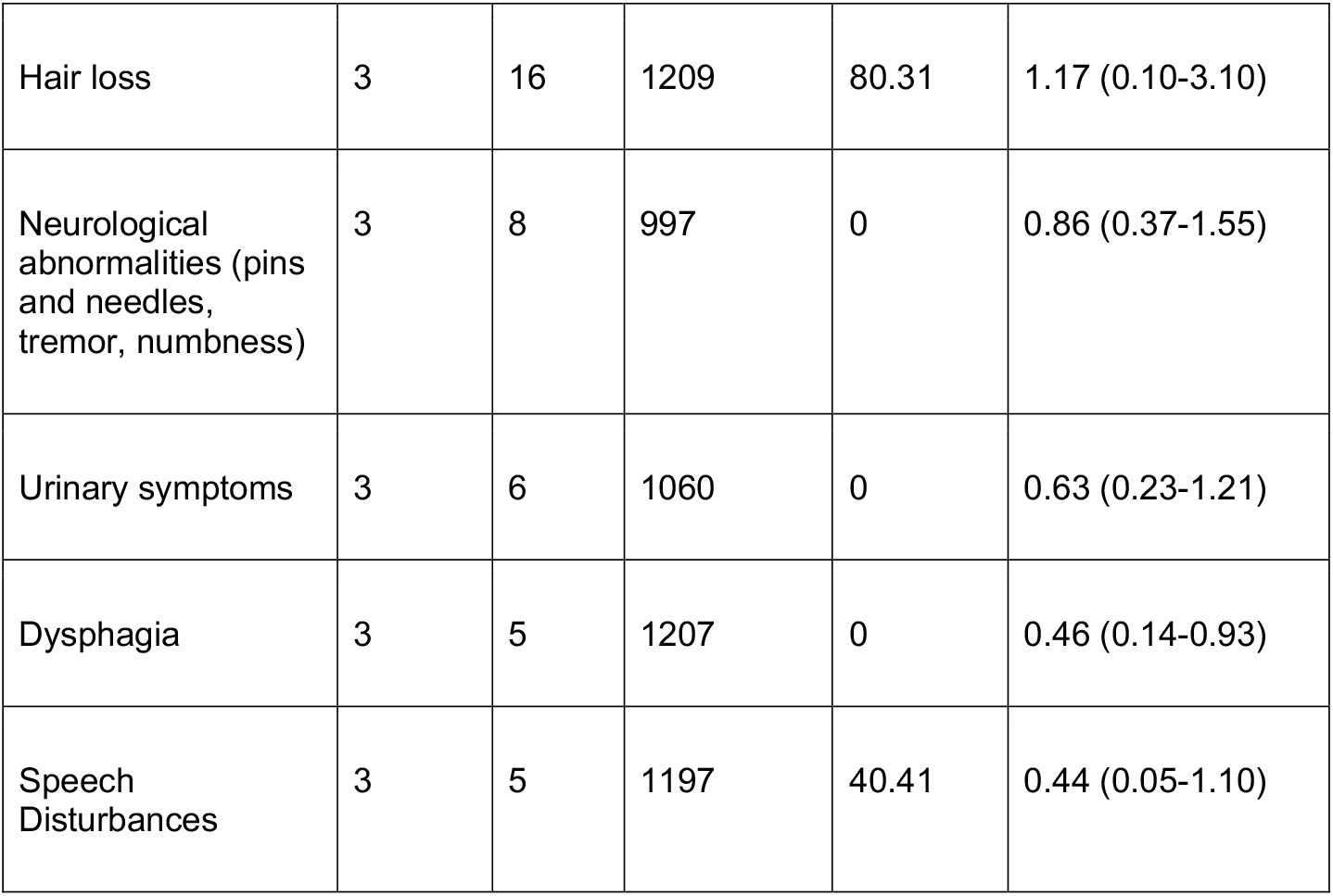
Pooled prevalence of symptoms in children and adolescents.

### Meta-analyses of the prevalence of long-COVID

The prevalence of long-COVID in minors, as defined by the presence of one or more symptoms more than 4 weeks following a SARS-CoV-2 infection, was 25.24% (95% CI, 18.17-33.02, I^2^ 99.61%) (Figure 2 and Figure 3). For hospitalized patients, the prevalence of long-COVID was 29.19% (95% CI, 17.83-41.98, I^2^ 80.84%). The most common symptoms and percentage of prevalence associated were mood symptoms (e.g., sadness, tension, anger, depression, and anxiety) (16.50%; 95% CI, 7.37-28.15, I^2^ 97.49%), fatigue (9.66%; 95% CI, 4.45-16.46, I^2^ 99.12%), sleep disorders (e.g., insomnia, hypersomnia, and poor sleep quality) (8.42%; 95% CI, 3.41-15.20, I^2^ 93.49%); headache (7.84%; 95% CI, 4.04-12.70, I^2^ 98.49%), respiratory symptoms (7.62%; 95% CI, 2.08-15.78, I^2^ 99.15%), sputum production or nasal congestion (7.53%; 95% CI, 3.78-12.36, I^2^ 0%), cognitive symptoms (e.g., less concentration, learning difficulties, confusion, and memory loss) (6.27%; 95% CI, 4.46-8.35, I^2^ 91.32%), loss of appetite (6.07%; 95% CI, 3.95-8.59, I^2^ 93.54%), exercise intolerance (5.73%; 95% CI, 0.00-19.38, I^2^ 87.77%), and altered smell (e.g., hyposmia, anosmia, hypersomnia, parosmia, and phantom smell) (5.60%; 95% CI, 3.13-8.69, I^2^ 97.11%). All other symptoms had less than 5.00% prevalence (Table 2, Figure 2, and Supplementary Figure 1).

**Figure 2.**
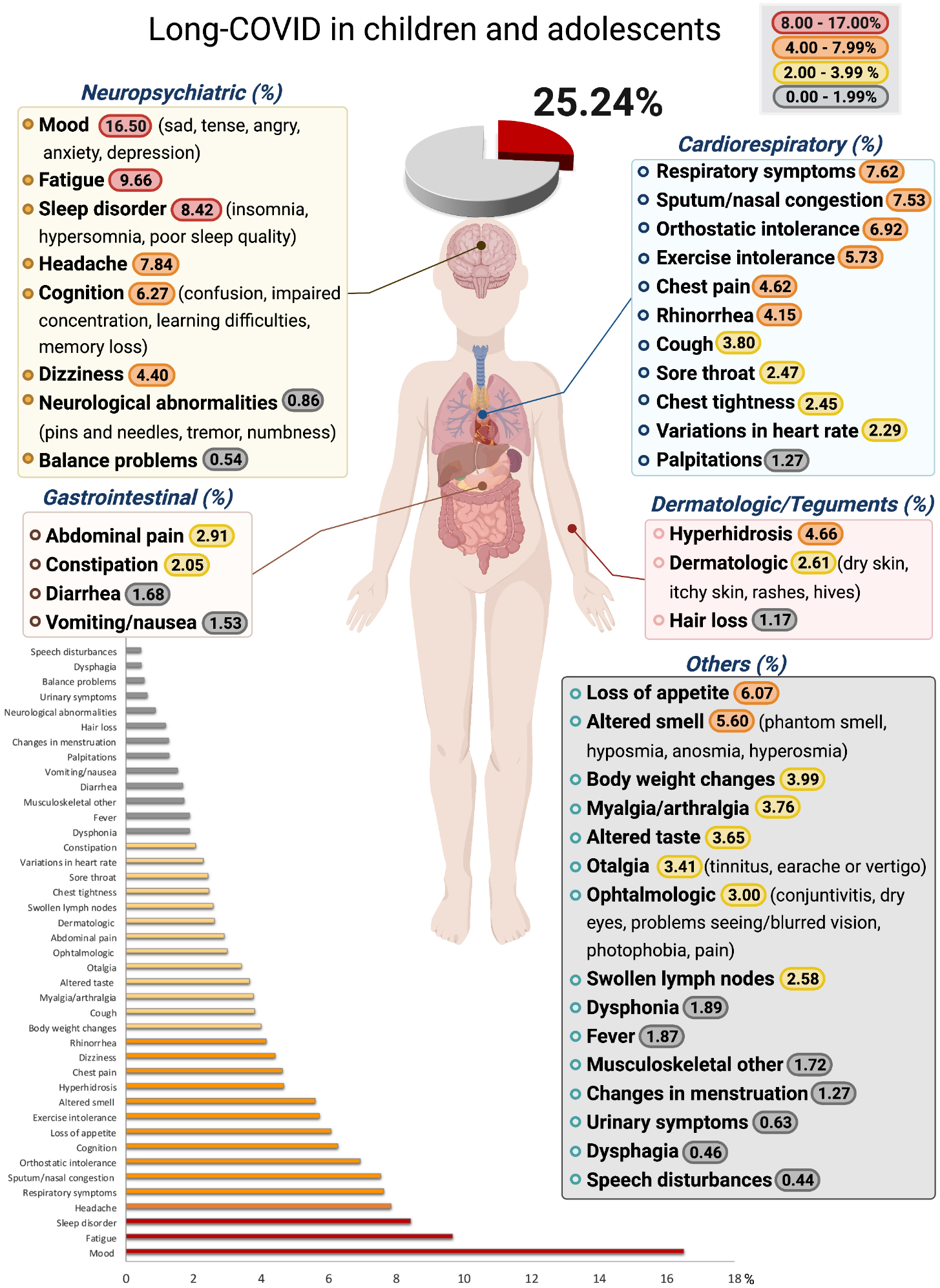
The pooled prevalence of long-COVID by symptoms in children and adolescents. Meta-analyses revealed that the prevalence of more than 40 long-COVID symptoms in children and adolescents. The presence of one or more symptoms following a SARS-CoV-2 infection was 25.24%.

**Figure 3.**
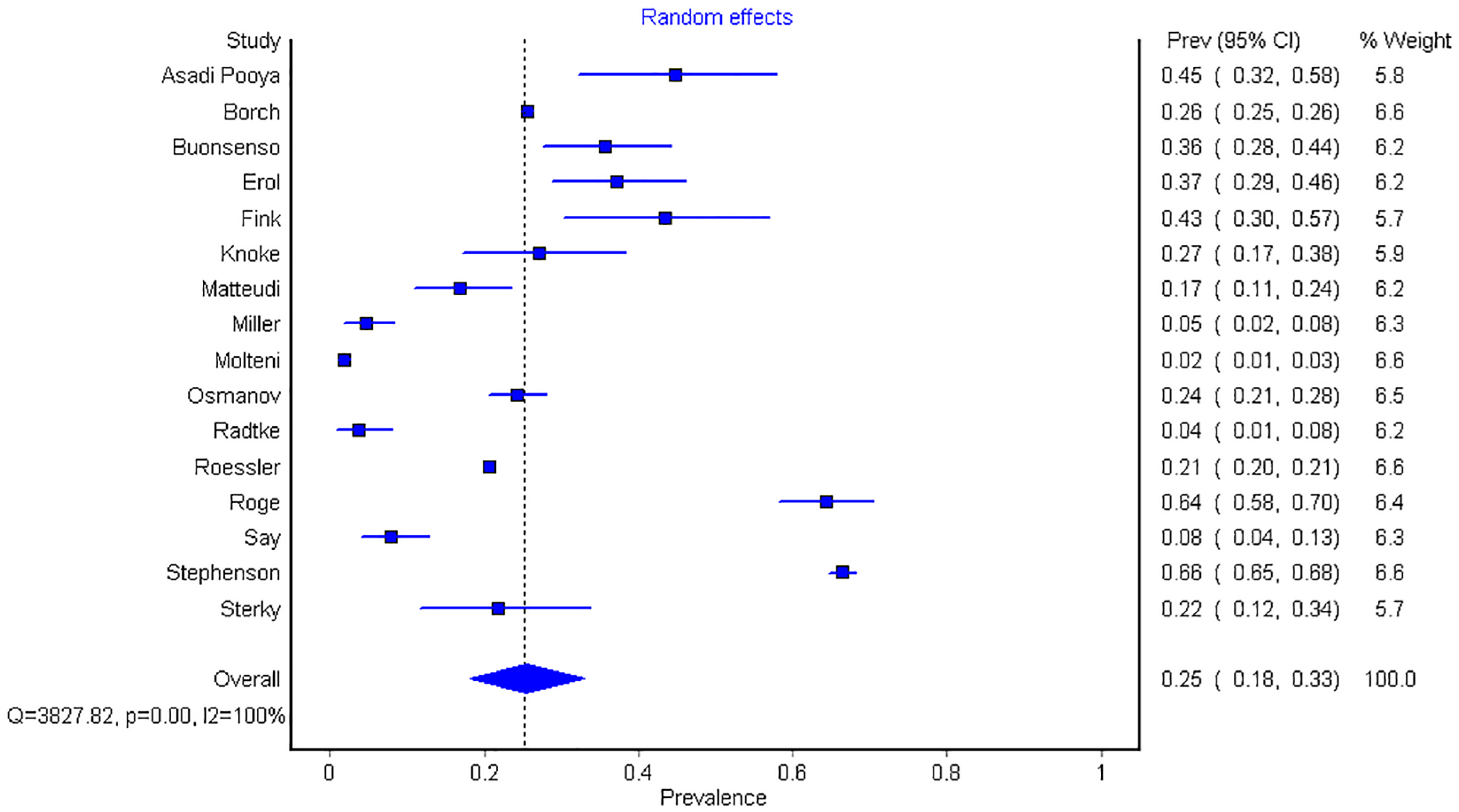
Forest Plot of pooled prevalence of long-COVID overall in children and adolescents.

### Meta-analyses of O.R.s (cases vs. controls)

It was only possible to perform meta-analyses of O.R.s comparing cases and controls for 13 symptoms (mood, fatigue, headache, dyspnea, concentration problems, anosmia/ageusia, loss of appetite, rhinitis, myalgia/arthralgia, cough, fever, sore throat, and nausea/vomiting) (Figure 4). When compared to controls, children with long-COVID had a higher risk of persistent dyspnea (OR:2.69; 95%CI, 2.30-3.14, I^2^ 0%), anosmia/ageusia (OR:10.68; 95% CI, 2.48, 46.03, I^2^ 0%), and/or fever (OR:2.23; 95% CI, 1.2-4.07, I^2^ 12%). There was significant heterogeneity for 4 out of the 13 meta-analyses (Figure 4). The controls were chosen in a very different way among studies, which might have introduced significant heterogeneity. The following were the different definitions of controls, minors with: 1) other infections (e.g., common cold, pharyngotonsillitis, gastrointestinal, urinary tract infections, pneumonia of bacteria or unknown origin)^17^; 2) no antibodies testing^24^ mixed with other infections^17^; 3) a negative antibody test^29^, 4) a negative rtPCR test among symptomatic minors^35^; and 5) minors who did not have a positive test recorded in the database^15^ (Supplementary Figure 2). The adjustments among studies also varied. Several studies adjusted their OR by age, sex, ethnicity, socioeconomic status, and comorbidities ^35^. However, age and sex^15^ only adjusted for sex, only for age^17^, did not adjust, or by OR without adjusting previous conditions^24^.

**Figure 4.**
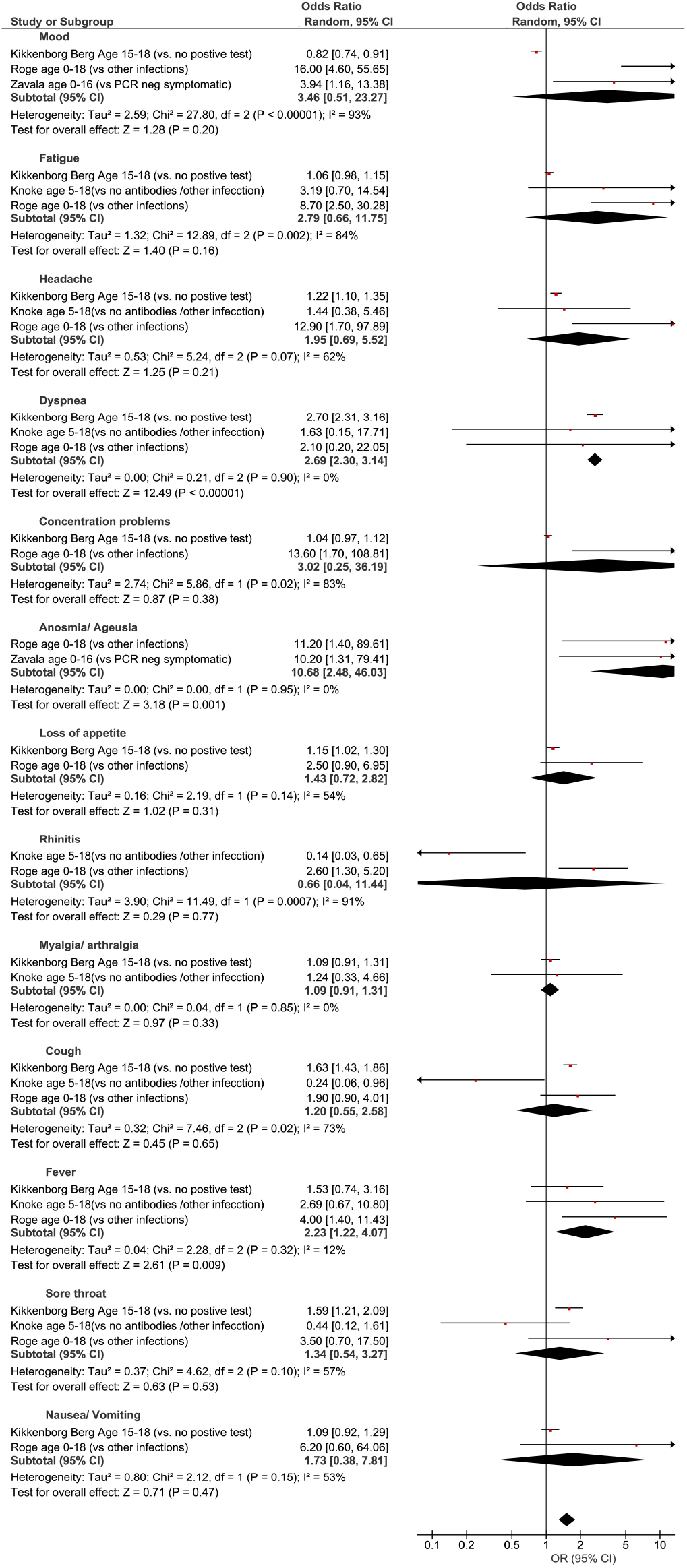
Pooled Odds Ratios with 95% CI in cases vs. controls. The size of each box indicates the effect of each study by symptom assigned using the Odds ratios (95% CI) by age and domain.

### Other Findings

The prevalence of symptoms over the course of long-COVID for cases and controls is shown in Supplementary Figure 2. Given the heterogeneity in the definition of controls and the low number of subjects, no formal statistical comparison was done for the crude prevalence. The symptoms that were presented in a single study and, therefore, unable to be incorporated into the meta-analyses, include: orthostatic intolerance, cold hands/feet, chapped lips, adenopathy, fainting, twitching of fingers and toes, chills, swollen toes/fingers, and hallucinations.

One study reported statistically significant differences between clinical cases and controls for systolic blood pressure, left ventricular ejection fraction, relative myocardial wall thickness, and tricuspid annular plane systolic excursion^22^. However, given that these variables were only evaluated in this study, we could not perform a meta-analysis for these outcomes. Studies included in the meta-analyses evaluated whether certain variables increased the risk of long-COVID and found that age, sex, severe acute-COVID-19, obesity, allergic disease, and long-term health conditions^19,26,28,36^. Further, two of the studies evaluated the duration of symptoms. A study from Denmark reported that symptoms resolved in a minimum of 54–75% of minors within 1–5 months^16^. Another from England, reported that 4.4% of children still had symptoms four weeks after COVID-19 onset, which decreased to 1.8% at 8 or more weeks^27^.

### Quality of studies

Regarding the quality of studies, all had a score of 7 or more (Supplementary Table 1). Table 3 presents a list of methodological strengths or limitations for each study. All studies included laboratory-confirmed COVID-19 infection, rt-PCR, antigen, or antibody test. Two-thirds of the studies included over 100 children. Six meta-analyses had low heterogeneity (I^2^<25%) for the following symptoms: vomiting and nausea, nasal congestion, dysphonia, urinary problems, neurological abnormalities, and dysphagia. Three meta-analyses had medium heterogeneity for the following symptoms: abdominal pain, changes in menstruation, and speech disturbances. All other meta-analyses had high heterogeneity (I^2^>75%). Given that very few studies by the different variables (e.g., age, sex, country, or past comorbidities), there were not enough studies that included this information to evaluate where the heterogeneity originated.

**Table 3.**
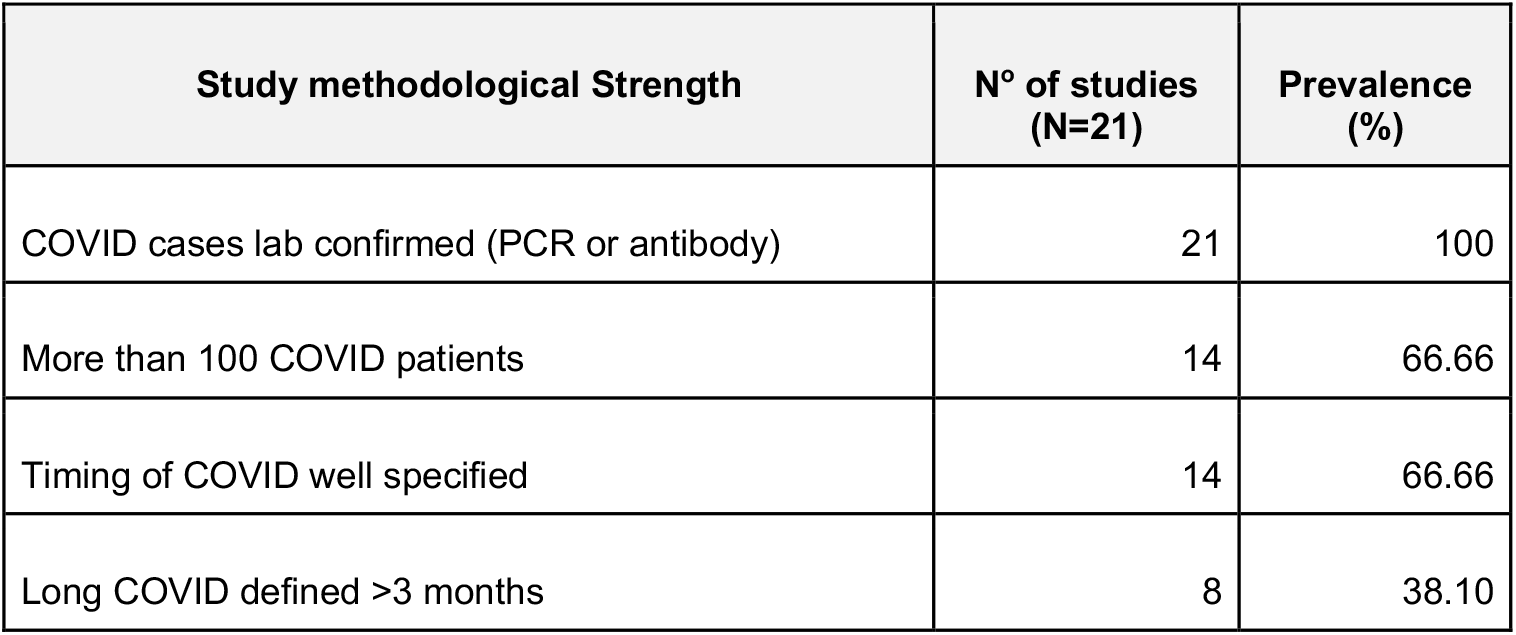
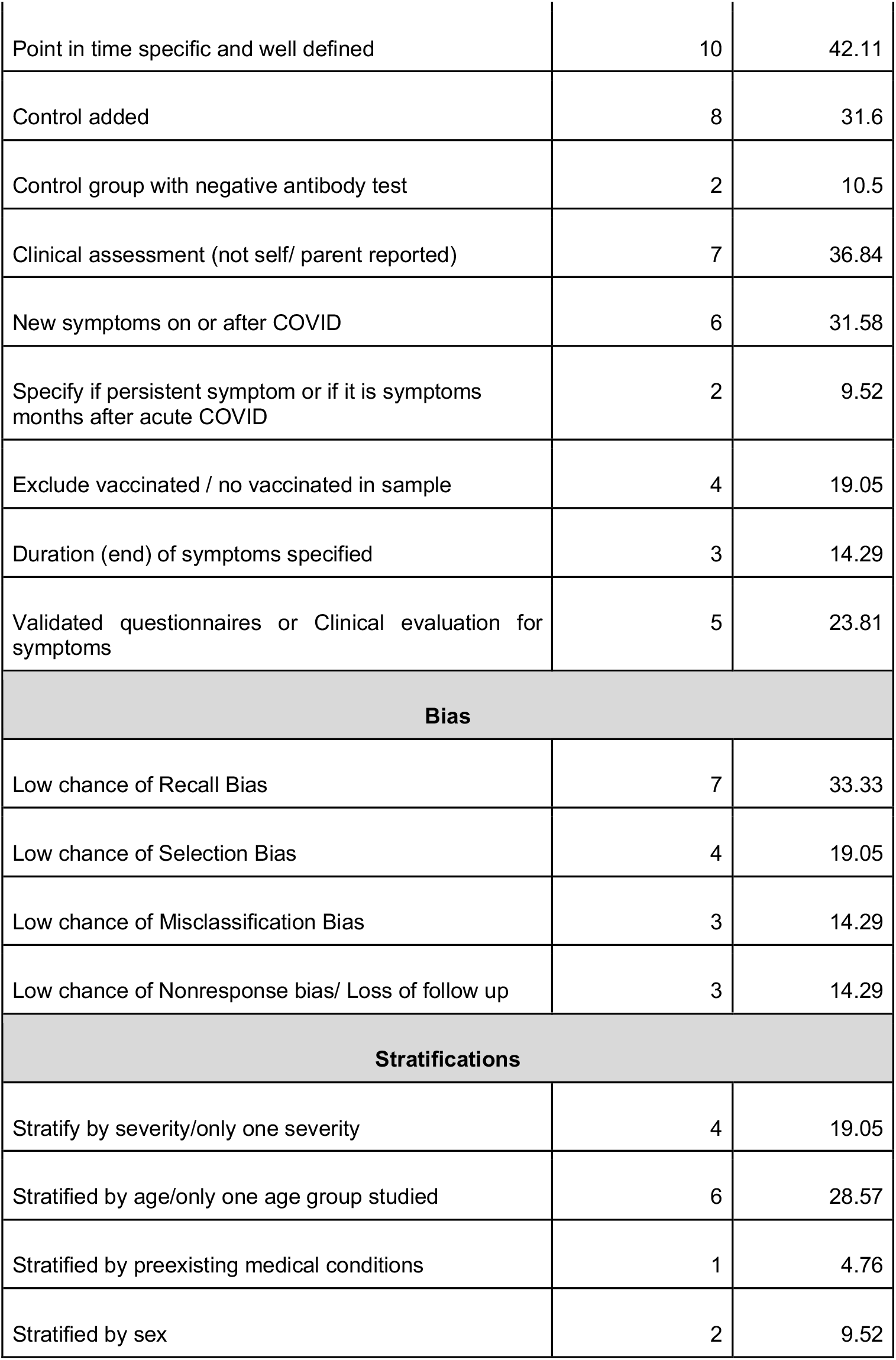
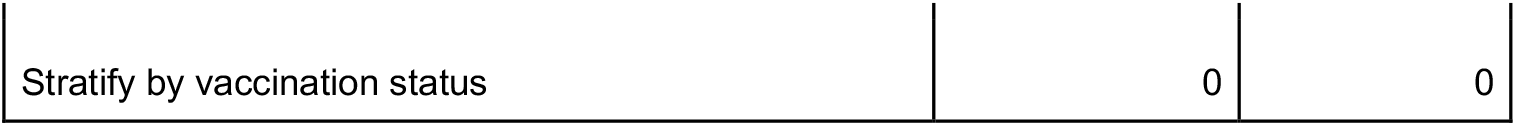
Study methodological strengths and limitations.

## DISCUSSION

The prevalence of long-COVID in children and adolescents was 25.24%. The five most prevalent clinical manifestations were mood symptoms (16.50%), fatigue (9.66%), sleep disorders (8.42%), headache (7.84%), and respiratory symptoms (7.62%). It was only possible to perform meta-analyses of ORs comparing cases and controls for 13 symptoms, with a higher risk of persistent dyspnea, anosmia/ageusia, and/or fever. Studies have shown that the pandemic has profoundly impacted society by affecting children’s development through isolation, poverty, food insecurity, loss of parents and caregivers, loss of time in education, and increased stress^37^. COVID-19 pandemic has initiated an explosion of future mental illnesses^38^, affecting both society as a whole and those who recover from long-COVID. The presence of these symptoms in the general population, regardless of COVID-19 status, has been coined long-Pandemic Syndrome^20^.

Interestingly, many of the symptoms identified in these meta-analyses associated to long-COVID, such as mood, fatigue, sleep disorders, orthostatic intolerance, decreased concentration, confusion, memory loss, balance problems, exercise intolerance, hyperhidrosis, blurred vision, body temperature dysregulation, dysfunction on heart, rate variability and palpitations, constipation or diarrhea, and dysphagia, are commonly present in dysautonomia^39^. Dysautonomia is a dysfunction of the sympathetic and/or parasympathetic autonomic nervous system. However, it remains unclear whether dysautonomia may be a direct result of the SARS-CoV-2 infection, interaction with other viruses, or immune-mediated processes such as cytokines, which are known mediators of the inflammatory response^40-43^. Moreover, the constellation of symptoms because of long-COVID can vary from patient to patient, fluctuating in their frequency and severity^44^. Like adults, the pediatric population’s risk factors associated with long-COVID are older age, female gender, severe COVID-19, overweight/obesity, comorbid allergic diseases, and other long-term co-morbidities. Protective factors leading to milder severity and duration of COVID-19, and possibly also long-COVID in children, include fewer comorbidities, strong innate immune responses, reduced expression of angiotensin-converting enzyme-2 (ACE2) receptors, and active thymic function, which leads to the increased presence and decreased depletion of T cells. Further protections include a range of environmental or non-inheritable factors such as vaccines, past infections, nutrition, and/or the gut microbiome^19,26,28,36,45^. Concerning age, most of the studies did not discriminate between children (<12-year-old) and adolescents. Given that the risk increases with age, there is a need for future studies to stratify by age. In addition, the evaluation of self-reporting symptoms can be significantly biased by age as younger children might not be able to express their emotional and functional status relevant to post-COVID-19 adequately. Further studies targeting age groups are required.

The prevalence of symptoms is highly dependent on how much time has passed after having acute COVID-19. The follow-up time in these meta-analyses varied between 1 to 13 months. Even though most symptoms improve with time ^46^, there is evidence in adult studies that suggests some symptoms can persist one year after COVID-19 diagnosis^47^. It is important to understand which symptoms are associated with certain periods of time (e.g., 6 months, 12 months, 2 years).

The strength of this meta-analyses focuses on the large sample size^48^ which helps identify the signs and symptoms present after acute SARS-CoV-2 infection. Further, there were some limitations such as the quality of the meta-analyses results depending on the quality of the studies included. Table 3 contains a list of all the methodological aspects that future studies need to consider. We can observe that all studies had a high probability of bias, including lack of standardized definitions, recall, selection, misclassification, nonresponse, and/or loss of follow-up. Additionally, the included studies have the limitations inherited in all observational studies, including bias due to residual and unmeasured confounding, and a high level of heterogeneity. To account for heterogeneity, we used a random-effects model^49^. The differences between studies were likely due to differences in study designs, settings, populations, follow-up time, symptom ascertainment methods, inconsistent terminology, little details on stratification of pre-existing comorbidities, and prior receipt of COVID-19 therapeutics and vaccines. Only four studies mentioned what percentage of the population was already vaccinated^15,16,20,28^ (Table 3). It has been shown that vaccines reduce the risk of long-COVID in adults ^50^. More studies are needed to analyze the relationship between vaccines in children with long COVID.

In our analyses, we included 2 pre-publication articles that are still not peer-reviewed ^26,30^. The limitations of the study of Roessler *et al*. ^*30*^ are inherited from all observational studies and are mentioned above. However its strengths are that it includes a large population (n=11,950), that physicians and psychotherapists validate the cases, and that they estimate the Incidence Rate Ratios (IRR), controlling for age, sex, and prevalent medical conditions by applying propensity score matching. They report that symptoms were statistically higher in the acute COVID-19 cohort (IRR=1.30) than in the controls without COVID-19. The study by Miller et al.^26^ included children participating in VirusWatch, a household cohort in England and Wales that recruited households via postcards, social media, and SMS that had as an objective to study acute COVID-19. Long-COVID was defined as answering “YES” to the following question - “*Have you presented any new symptoms that have lasted for four or more weeks even if these symptoms come and go and cannot be explained by something else?”*. Given the nature of the survey, it has a possibility of introducing bias to the results (recall bias, selection bias, misclassification bias, and nonresponse bias). The study does not present the frequencies for each symptom, and therefore it was not included in the meta-analyses of each symptom. We only included this study in the meta-analysis of overall long-COVID. The authors estimated a prevalence of 4.6%, which, compared to the other studies, is underestimated.

Future prospective studies should include a control cohort and stratify and/or adjust their results by age, sex, race, severity of acute COVID-19 infection paired with clinical evaluation, vaccination status, preexisting medical conditions, and, if possible, SARS-CoV-2 variant. If we had analyzed these types of factors separately, we would have been able to determine the variations in the prevalence of long-COVID. Retrospective studies using large population-based databases with historical controls and secondary data sources (e.g., claims and medical records) should also be used. The selection of controls will be difficult in the future because not all the cases are recorded in databases (e.g., home tests), tests can be false, negative, or positive, or children can be asymptomatic. Future studies’ proposed control groups include a negative N protein antibody test without vaccination, a negative antibody test with vaccination, or historical cohorts that include children who have neither been vaccinated nor exposed to the virus.

## CONCLUSION

Protective measures are essential to prevent long-COVID in children. We need to understand the long-COVID pathophysiology and symptomatology to support clinical management systems, establish rehabilitation programs, and design guidelines and therapeutic research. Long-COVID represents a significant public health concern, and there are no guidelines to address its diagnosis and management. Our meta-analyses further support the importance of continuously monitoring the impact of long-COVID in minors and the need to include all variables and appropriate control cohorts in studies to better understand the real burden of pediatric long-COVID better.

## Data Availability

All data relevant to the study are included in the article or uploaded as supplementary information. In addition, the datasets used and/or analyzed during the current study are available from the corresponding author upon reasonable request.

## Abbreviations

CI: Credible interval
COVID-19: Coronavirus disease 2019
CSS: cross-sectional study
DM: diabetes mellitus
QCed: Quality-Controlled
IRR: Incidence Rate Ratios
NICE: National Institute for Health and Care Excellence
MIS-C: multisystem inflammatory syndrome
ORs: Odds Ratios
PCS: prospective cohort study
PRISMA: Preferred Items for Systematic Reviews and Meta-Analyses
RCS: retrospective cohort study
rt-PCR: real-time reverse transcription-polymerase chain reaction
SARS-CoV-2: severe acute respiratory syndrome coronavirus 2
PASC: post-acute sequelae of SARS-CoV-2

## Acknowledgments

Figure 2 was created by S.V. using Biorender.com. The authors are indebted to Dr. Gillian Hamilton for editing.

## Funding

This article was funded by Houston Methodist Research Institute (S.V.).

## Contributions

A.C., C.A., P.R., and R.S. extracted data. S.L.L. performed the statistical analyses, T.W.O. performed the QC of the results. T.W.O., C.P., R.S., P.R., and S.V. performed the literature search, collected the data, wrote the manuscript, and made edits. S.L.L. and S.V. were mainly responsible for interpreting the data and preparing the final version. S.V. created the figures. All authors provided critical feedback and contributed to the final manuscript. Correspondence and requests for materials should be addressed to S.V.

## Ethics declarations

### Competing interests

The authors are solely responsible for all content, and funders played no role in study design, data collection and analysis, the decision to publish, or the preparation of the manuscript. S.L.L. is an employee of Novartis Pharmaceutical Company; the statements presented in the paper do not necessarily represent the position of the company. The remaining authors have no competing interests to declare.

## Notes

### Competing Interest Statement

SLL is an employee of Novartis Pharmaceutical Company; the statements presented in the paper do not necessarily represent the position of the company. The remaining authors have no competing interests to declare

### Funding Statement

This work was supported by funds from Houston Methodist Research Institute, Houston, TX.

### Summary of Updates

We have edited the text and minor changes to the figures.

