## Supplementary figures and images for "Long-COVID in Children and Adolescents: A Systematic Review and Meta-analyses"

### Supplemental Figure 1

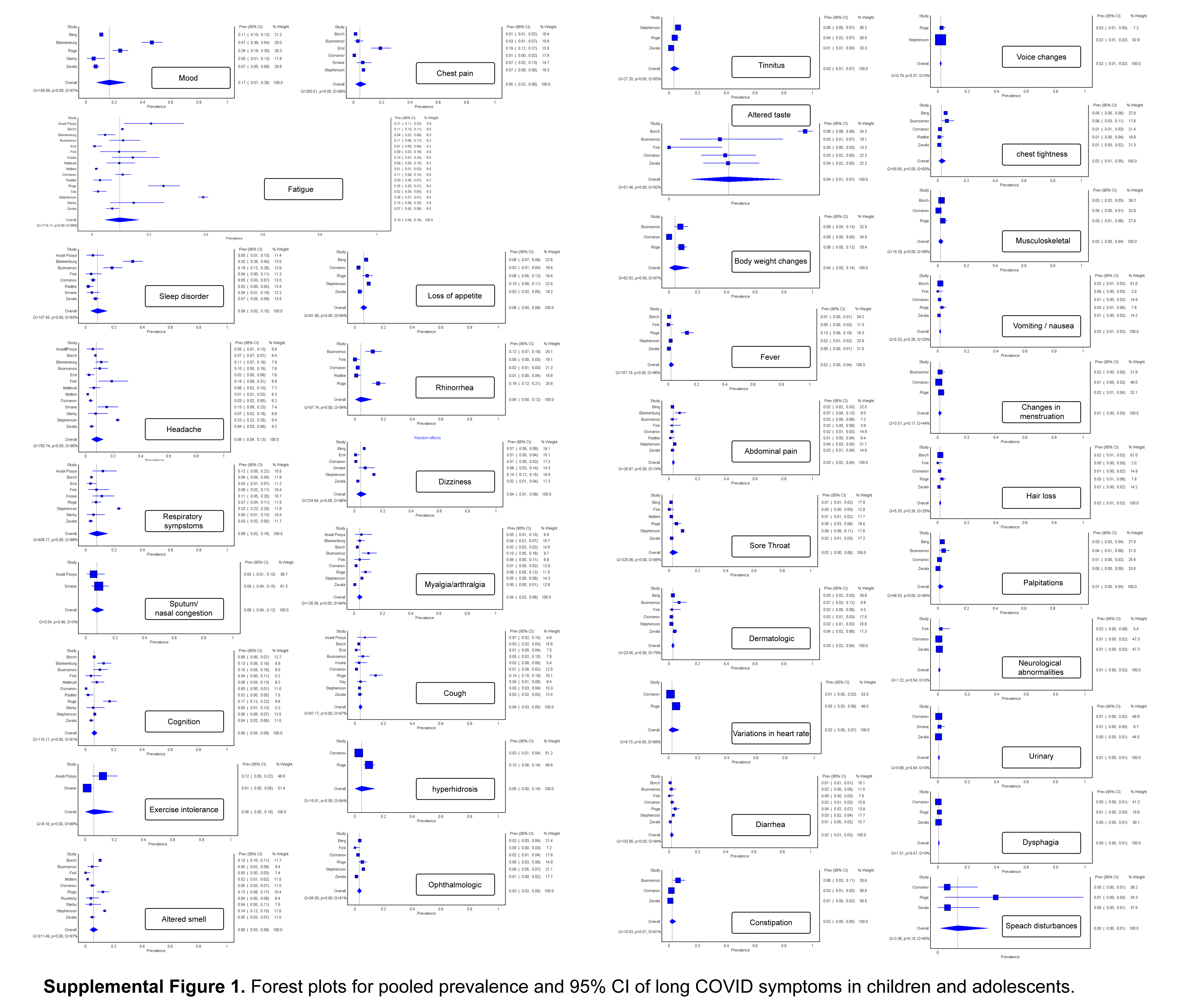

### Supplemental Figure 2

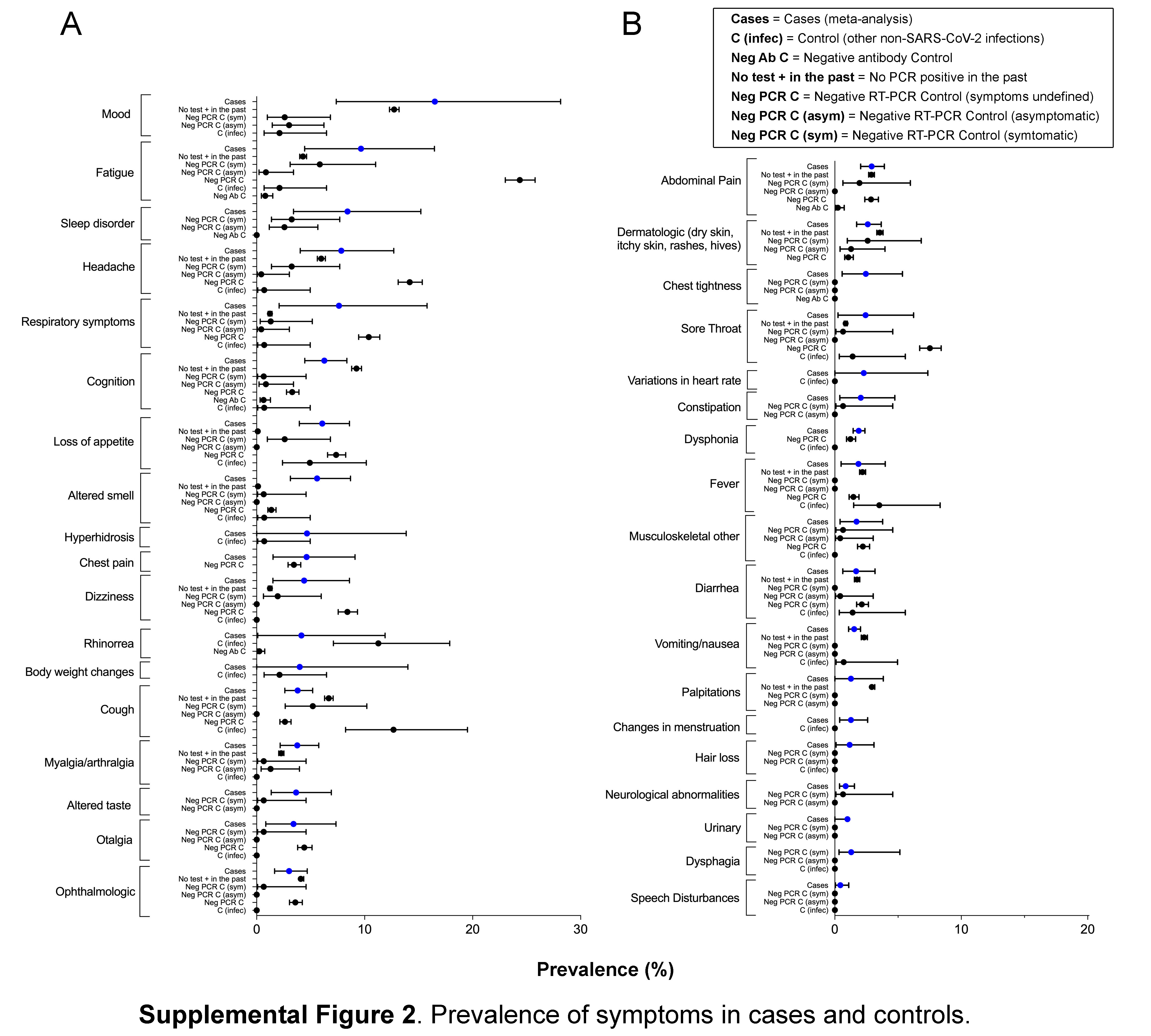
