## Supplemental Table 1 for "Long-COVID in Children and Adolescents: A Systematic Review and Meta-analyses"

**Supplemental Table 1.** Health states Quality Index variables.

| Study | 1. Population and observation period well defined | 2. Diagnostic criteria | 3. Method of case ascertainment | 4. Administration of measurement protocol | 5. Catchment Area | 6. Prevalence measure | Total (Max: 11) |
| --- | --- | --- | --- | --- | --- | --- | --- |
| Asadi-Pooya ^19^ | 1 | 0 | 1 | 3 | 2 | 2 | 9 |
| Blankenburg ^20^ | 0 | 1 | 2 | 3 | 1 | 2 | 9 |
| Borch ^16^ | 1 | 0 | 2 | 3 | 2 | 2 | 10 |
| Buonsenso ^21^ | 1 | 1 | 1 | 3 | 2 | 1 | 9 |
| Erol ^22^ | 0 | 1 | 1 | 3 | 1 | 1 | 7 |
| Fink ^23^ | 0 | 1 | 1 | 3 | 1 | 2 | 8 |
| Kikkenborg Berg* ^15^ | 1 | 1 | 2 | 3 | 2 | 2 | 11 |
| Knoke ^24^ | 1 | 1 | 2 | 3 | 2 | 2 | 11 |
| Matteudi ^25^ | 1 | 0 | 2 | 3 | 2 | 2 | 10 |
| Miller ^26^ | 0 | 1 | 2 | 3 | 1 | 2 | 9 |
| Molteni ^27^ | 0 | 0 | 2 | 3 | 1 | 2 | 8 |
| Osmanov ^28^ | 1 | 1 | 2 | 3 | 1 | 2 | 10 |
| Radtke ^29^ | 1 | 1 | 2 | 3 | 2 | 2 | 11 |
| Roge ^17^ | 0 | 0 | 2 | 3 | 2 | 2 | 9 |
| Roessler ^30^ | 1 | 1 | 2 | 3 | 2 | 2 | 11 |
| Rusetsky ^31^ | 1 | 1 | 1 | 3 | 1 | 1 | 8 |
| Say ^32^ | 1 | 1 | 1 | 3 | 1 | 2 | 9 |
| Smane ^18^ | 1 | 1 | 1 | 3 | 1 | 2 | 9 |
| Stephenson ^33^ | 0 | 1 | 2 | 3 | 1 | 2 | 9 |
| Sterky ^34^ | 1 | 1 | 1 | 3 | 1 | 2 | 9 |
| Zavala ^35^ | 0 | 1 | 2 | 3 | 2 | 2 | 10 |

1) Yes=1, No=0

2) Diagnostic system reported=1, Own system /symptoms described/no system/not specified = 0;

3) Community survey/multiple institutions=2, Inpatient/inpatients, and outpatients/case registers=1, Not specified=0

4) Administered interview=3, Systematic case note review=2, Chart diagnosis/case records=1, Not specified=0

5) Broadly representative (national or multi-site survey) =2, Small area/not representative (single community, single university) =1, Convenience sampling/ other (primary care sample/treatment group) =0

6) Point prevalence (e.g. one month) =2, 12-month prevalence=1, Lifetime prevalence = 0
